# Combined point of care SARS-CoV-2 nucleic acid and antibody testing in suspected moderate to severe COVID-19 disease

**DOI:** 10.1101/2020.06.16.20133157

**Authors:** Petra Mlcochova, Dami Collier, Allyson Ritchie, Sonny M. Assennato, Myra Hosmillo, Neha Goel, Bo Meng, Krishna Chatterjee, Vivien Mendoza, Nigel Temperton, Leo Kiss, Leo C. James, Katarzyna A. Ciazynska, Xiaoli Xiong, John AG Briggs, James Nathan, Federica Mescia, Hongyi Zhang, Petros Barmpounakis, Nikos Demeris, Richard Skells, Paul A. Lyons, John Bradley, Steven Baker, Jean Pierre Allain, Kenneth GC Smith, Ian Goodfellow, Ravindra K. Gupta

## Abstract

**Background:** Rapid COVID-19 diagnosis in hospital is essential for patient management and identification of infectious patients to limit the potential for nosocomial transmission. The diagnosis of infection is complicated by 30-50% of COVID-19 hospital admissions with nose/throat swabs testing negative for SARS-CoV-2 nucleic acid, frequently after the first week of illness when SARS-CoV-2 antibody responses become detectable. We assessed the diagnostic accuracy of combined rapid antibody point of care (POC) and nucleic acid assays for suspected COVID-19 disease in the emergency department.

**Methods:** We developed (i) an in vitro neutralization assay using a lentivirus expressing a genome encoding luciferase and pseudotyped with spike (S) protein and (ii) an ELISA test to detect IgG antibodies to nucleocapsid (N) and S proteins from SARS-CoV-2. We tested two lateral flow rapid fingerprick tests with bands for IgG and IgM. We then prospectively recruited participants with suspected moderate to severe COVID-19 and tested for SARS-CoV-2 nucleic acid in a combined nasal/throat swab using the standard laboratory RT-PCR and a validated rapid POC nucleic acid amplification (NAAT) test. Additionally, serum collected at admission was retrospectively tested by *in vitro* neutralisation, ELISA and the candidate POC antibody tests. We evaluated the performance of the individual and combined rapid POC diagnostic tests against a composite reference standard of neutralisation and standard laboratory based RT-PCR.

**Results:** 45 participants had specimens tested for nucleic acid in nose/throat swabs as well as stored sera for antibodies. Using the composite reference standard, prevalence of COVID-19 disease was 53.3% (24/45). Median age was 73.5 (IQR 54.0-86.5) years in those with COVID-19 disease by our composite reference standard and 63.0 (IQR 41.0-72.0) years in those without disease. The overall detection rate against the composite reference standard was 79.2% (95CI 57.8-92.9%) for rapid NAAT, decreasing from 100% (95% CI 65.3-98.6%) in days 1-4 to 50.0% (95% CI 11.8-88.2) for days 9-28 post symptom onset. Correct identification of COVID-19 with combined rapid POC diagnostic tests was 100% (95CI 85.8-100%) with a false positive rate of 5.3-14.3%, driven by POC LFA antibody tests.

**Conclusions:** Combined POC tests have the potential to transform our management of COVID-19, including inflammatory manifestations later in disease where nucleic acid test results are negative. A rapid combined approach will also aid recruitment into clinical trials and in prescribing therapeutics, particularly where potentially harmful immune modulators (including steroids) are used.

## Introduction

As of the 22^nd^ of June 2020, 9.0 million people have been infected with SARS-CoV-2 with over 469,939 deaths(Dong et al., 2020). The unprecedented numbers requiring SARS-CoV-2 testing has strained healthcare systems globally. There is currently no gold standard for diagnosis of COVID-19. Detection of SARS-CoV-2 by nucleic acid amplification testing (NAAT), is largely done by real time RT-PCR on nose/throat swabs in centralised laboratories. RT-PCR specimens need to be handled in containment level 3 category laboratory (CL3) and then batch analysed. Given these bottlenecks, the turnaround time for this test is in the order of 2-4 days(Collier et al., 2020). NAAT tests from a single nose/throat swab are negative in up to 50% in patients who have CT changes consistent with COVID-19 and/or positive antibodies to SARS-CoV-2 (Arevalo-Rodriguez et al., 2020; Fang et al., 2020; Wang et al., 2020b). The lack of detectable virus in upper airway samples is not only a serious barrier to making timely and safe decisions in the ER, but also leads to multiple swab samples being sent, frequently from the same anatomical site, leading to additional strain on virology laboratories. Nonetheless, NAAT remains important in identifying infectious individuals. Additionally, in severely ill patients tracheo-bronchial samples might be NAAT positive even when the nose/throat swab is negative(Tang et al., 2020; Wang et al., 2020b).

Multiple factors might contribute to negative results by NAAT, including test sensitivity, sampling technique and timing of the sampling in the disease course(Tang et al., 2020). The viral load in the upper respiratory tract is detectable from around 4 days before symptoms(Arons et al., 2020) and frequently wanes after a week post symptom onset(He et al., 2020) (Lescure et al., 2020). Similarly, a case series from Germany found the detection rate by RT-PCR was <50% after 5 days since onset of illness(Wolfel et al., 2020). A proportion of patients develop a secondary deterioration in clinical condition requiring hospitalisation and respiratory support, at a time when immune pathology is thought to be dominant rather than direct pathology related to viral replication (Lescure et al., 2020; Siddiqi and Mehra, 2020).

An antibody response to SARS-CoV-2 is detectable 6 days from infection and is almost always neutralising (Long et al., 2020; Suthar et al., 2020). Antibody based diagnosis of COVID-19 shows increasing sensitivity in the latter part of the infection course when NAAT testing on nose/throat samples is more likely to be negative(Lassaunière et al., 2020; Liu et al., 2020; Pickering et al., 2020; Whitman et al., 2020). As a result, diagnosis of infection as well as identification of infectivity would benefit from a combination of virologic and immunologic markers to inform patient initial triage and subsequent management. It is critical to determine whether a rapid point of care combined antibody and nucleic acid testing strategy could improve diagnosis.

We previously evaluated the diagnostic accuracy of the SAMBA II SARS-CoV-2 rapid test compared with the standard laboratory RT-PCR and found similar accuracy with a turnaround time of 2-3 hours even in real world settings (Collier et al., 2020). Several studies have now reported head-to-head comparisons of immuno-chromatographic lateral flow immunoassays (LFAs)(Adams et al., 2020; Lassaunière et al., 2020; Pickering et al., 2020; Whitman et al., 2020). These assays are cheap to manufacture and give a binary positive/negative result, thereby lending themselves well to point of care (POC) testing. Even though they have variable performance and in general are negative in the early phase of infection, they become highly sensitive in the later stage of illness(Adams et al., 2020; Lassaunière et al., 2020; Pickering et al., 2020; Whitman et al., 2020). In this study we evaluated the diagnostic performance of a POC combination comprising NAAT and LFA antibody testing against a composite reference standard of laboratory RT-PCR and a serum neutralisation assay.

## Results

45 prospectively recruited participants with suspected moderate to severe COVID-19 disease had specimens tested for nucleic acid in nose/throat swabs as well as stored sera for antibody testing. Samples at hospital admission were collected at a median of 7 (IQR 7-13) days after illness onset. Results from the four IgG antibody assays utilised in this study were confirmed (4 or 3 concordant) in 38/45 samples and, against this classification, neutralisation, spike ELISA ((Amanat et al., 2020) and Supplementary Figure 1), Surescreen and COVIDIX Healthcare assays (Figure 1C) gave a correct result in 100%, 97.4%, 92.1% and 86.8%, respectively, justifying the choice of the neutralisation assay as standard.

**Figure 1:**
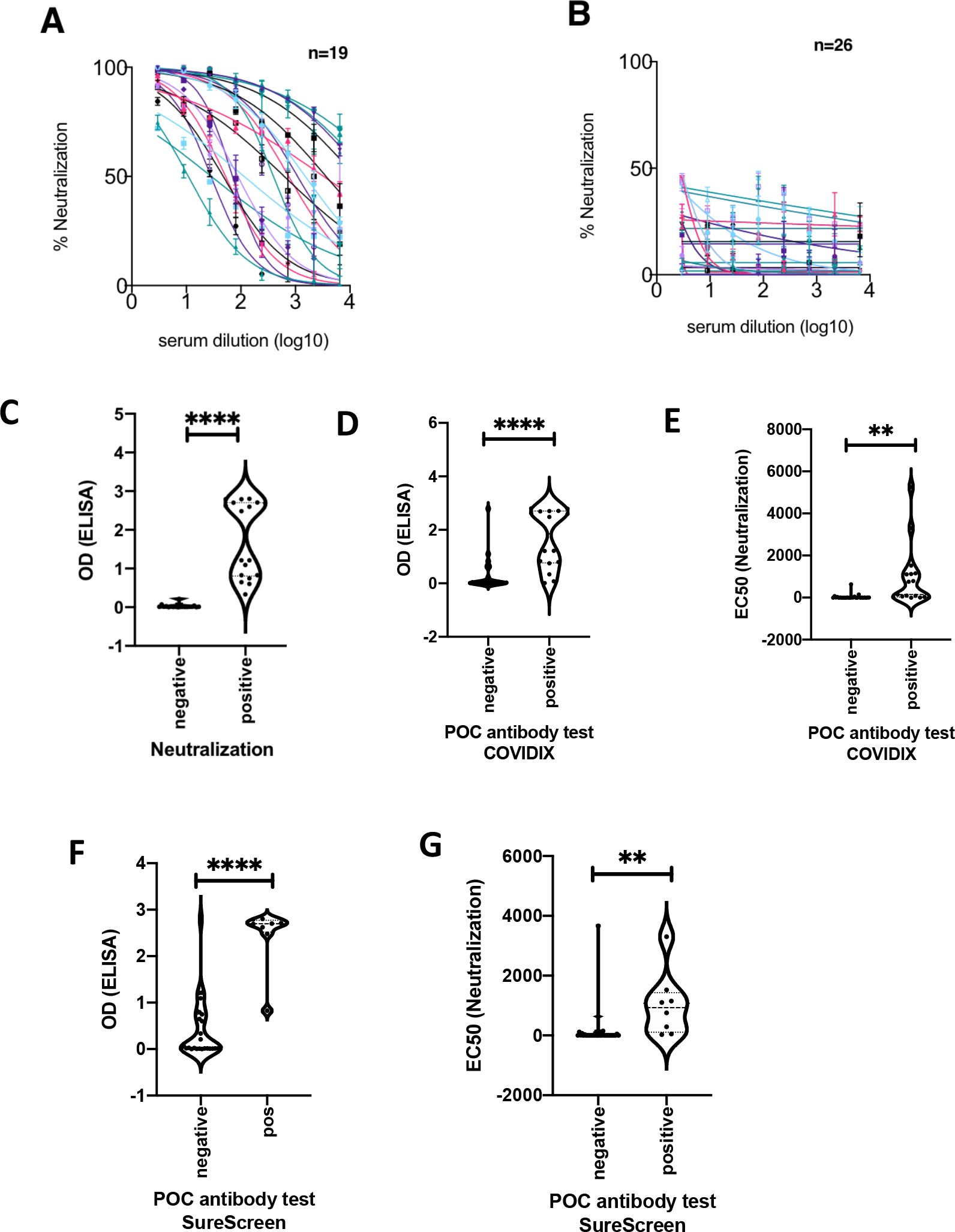
Antibody detection for SARS-CoV-2: cross validation of lateral flow diagnostic tests (POC antibody) with ELISA and SARS-CoV-2 pseudotype virus neutralisation assays. A, B. Serum from COVID-19 suspected participants inhibited (n=19) (A) or did not inhibit (n=26) (B) SARS-CoV-2 pseudotype virus infection in a neutralisation assay. Serum from a healthy donor was used and a negative control. The assay was performed in duplicate. Error bars represent SEM. C. Comparison between ELISA and positive/negative results from neutralisation assay. n=37, p<0.0001. D. Comparison between ELISA Spike protein reactivity and positive/negative POC antibody test results (COVIDIX SARS-CoV-2 IgM/IgG Test). n=38, p<0.0001. E. Comparison between EC50 dilution titre from neutralisation assay and positive/negative POC antibody test results (COVIDIX SARS-CoV-2 IgM/IgG Test). n=44, p=0.0025. F. Comparison between ELISA IgG and positive/negative POC IgG band results for SureScreen SARS-CoV-2 IgM/IgG test. n=38, p<0.0001. G. Comparison between EC50 dilution titre from neutralisation assay and positive/negative SureScreen SARS-CoV-2 IgM/IgG antibody band test results. n=43, p=0.005.

The sera from 42.2% (19/45) participants showed strong neutralising antibody response against SARS-CoV-2 spike protein pseudotyped virus infection in a neutralization assay (Figure 1A). 26 participants’ sera showed no neutralising response (Figure 1B). The neutralisation ability of participants’ sera was compared with an in house ELISA IgG assay for Spike specific antibodies based on a recently reported method(Amanat et al., 2020) (Supplementary Figure 1), and significant association between positive results in both assays was demonstrated (Figures 1C, p<0.0001). Figures 1D-G show significant associations between the point of care antibody test result and both ELISA (p<0.0001) and neutralisation assays, p<0.0025. Importantly, the neutralisation assay also confirmed no cross-reactivity of test sera with SARS-CoV-1 (Supplementary Figure 2).

53.3% (24/45) of participants had COVID-19 disease, as determined by the composite reference standard (lab RT-PCR and neutralisation assay). Median age was 73.5 (IQR 54.0-86.5) years in those with SARS-CoV-2 infection by our composite reference standard and 63.0 (IQR 41.0-72.0) years in those without disease (Table1). CRP and procalcitonin were significantly higher in confirmed COVID-19 patients and ‘classical’ chest radiograph appearances were more common in confirmed COVID-19 patients (Table1, p<0.001). However, 6/24 (25%) had normal or indeterminate chest radiographs in the confirmed COVID-19 group. 14/24 (58.3%) patients deemed to be COVID-19 positive by the reference composite standard were positive by both rapid NAAT and antibody testing.

The overall COVID-19 diagnosis rate (positive predictive agreement) by rapid nucleic acid testing was 79.2% (95% CI 57.8-92.9), decreasing from 100% (95% CI 65.3-98.6%) for days 1-4 to 50.0% (95% CI 11.8-88.2) for days 9-28 post symptom onset (Table 2 and Supplementary Figure 3). When IgG/IgM rapid tests were combined with NAAT, the overall positive predictive agreement increased to 100% (95% CI 85.8-100);100% (95% CI 59.0-100) in days 1-4 of illness and 100% (95% CI 54.1-100) in days 9-28 of illness for both POC antibody tests (Table 2). However, among 21 COVID-19 negative individuals, there were three false positive results for one POC antibody test and one false positive result for the other, resulting in positive predictive values of 88.9% and 96.0% for the two POC antibody/ SAMBA II NAAT combinations (Table 2). On closer analysis of ‘false positive’ results for the POC tests, we noted that two individuals had normal chest radiographs and the third had a pulmonary embolus diagnosed on CT pulmonary angiography. All had normal lymphocyte counts (Supplementary table 1).

**Table 1:** Characteristics of participants in prospective study. COVID-19 status is based on composite gold standard test of nose/throat swab SARS-CoV-2 RT-PCR + serum neutralisation of pseudovirus bearing SARS-CoV-2 Spike. ^§^ Wilcoxon rank sum test used except where indicated. ^a^ Chi-square test.

|  | COVID-19<br>N=24 | No COVID-19<br>N=21 | P value<sup>§</sup> |
| --- | --- | --- | --- |
| Male sex (%) | 14 (58.3) | 9 (42.9) | 0.30<sup>a</sup> |
| Median age (IQR) yrs | 73.5 (54.0-86.5) | 63.0 (41.0-72.0) | 0.03 |
| Median SpO2 (IQR) % | 95.0 (92.5-96.0) | 96.0 (94.0-98.0) | 0.09 |
| Median FiO2 (IQR) | 0.21 (0.21-0.24) | 0.21 (0.21-0.21) | 0.40 |
| Median PaO2 (IQR) Kpa | 5.0 (3.0-9.1) | 7.2 (3.8-9.0) | 0.30 |
| Median PaO2:FiO2 ratio (IQR) | 20.5 (13.3-32.9) | 30.9 (18.1-36.2) | 0.09 |
| Median Respiratory rate (IQR) breaths/min | 22.0 (19.0-27.5) | 20.0 (17.0-23.0) | 0.06 |
| Median heart rate (IQR) beats/min | 86.0 (77.5-99.5) | 88.0 (78.0- 107.0) | 0.44 |
| Median Systolic BP (IQR) mmHg | 139.5 (117.5-149.0) | 135.0 (119.0-152.0) | 0.90 |
| Median duration of illness (IQR) days | 7 (1-8) | 10 (3-14) | 0.10 |
| Median Hb (IQR) g/dL | 12.9 (12.0-13.8) | 13.1 (11.6-14.1) | 0.46 |
| Median WCC (IQR) x10<sup>9</sup>/L | 7.0 (5.0-8.0) | 9.0 (7.0-14.0) | 0.08 |
| Median lymphocyte count (IQR) x10<sup>9</sup>/L | 0.8 (0.5-1.2) | 1.2 (0.8-1.5) | 0.12 |
| Median platelet count (IQR) x10<sup>9</sup>/L | 213.5 (188.5-303.5) | 271.0 (186.0-305.0) | 0.59 |
| Median Ferritin (IQR) µg/L | 684.7 (206.2-1059.1) | 112.3 (49.6-323.6) | 0.02 |
| Median Dimer (IQR) ng/mL | 369.0 (254.0-974.0) | 267.5 (66.0-550.5) | 0.10 |
| Median CRP (IQR) mg/L | 72.0 (28.5-214.5) | 12 (4.0-53.0) | 0.004 |
| Median procalcitonin (IQR) ng/mL | 0.2 (0.1-0.6) | 0.0 (0.0-0.1) | 0.03 |
| Radiological findings |  |  | <0.001<sup>a</sup> |
| Normal | 2 (8.3) | 9 (42.9) |  |
| Indeterminate | 4 (16.7) | 3 (14.3) |  |
| Classic | 18 (75.0) | 3 (14.3) |  |
| Non-COVID | 0 (0.0) | 6 (28.5) |  |

**Table 2.** Individual and combined diagnostic accuracy of point of care rapid NAAT-based and antibody tests according to time from initial symptoms. Positivity predictive agreement is the percentage of positive test results in samples deemed positive by the composite reference standard. Negative predictive agreement is the percentage of negative test results in samples deemed negative by the composite reference standard. *43 out of 45 patients had SureScreen antibody results

| % (95% CI) | Days 1-4<br>N=14 | Days 5-8<br>N=14 | Days 9-28<br>N=17 | Overall<br>N=45* |
| --- | --- | --- | --- | --- |
| **SAMBA II SARS-CoV-2**<br>Positive predictive agreement<br>Negative predictive agreement | 100 (65.3-98.6)<br>100 (69.2-100) | 81.8 (48.2-97.8)<br>100 (29.2-100) | 50.0 (11.8-88.2)<br>100 (71.5-100) | 79.2 (57.8-92.9)<br>100 (83.9-100) |
| **COVIDIX Ig M & IgG**<br>Positive predictive agreement<br>Negative predictive agreement | 100 (59.0-100)<br>100 (59.0-100) | 90.9 (58.7-99.8)<br>66.7 (9.4-99.2) | 100 (54.1-100)<br>81.8 (48.2-97.7) | 95.8 (78.9-99.9)<br>85.7 (63.7-97.0) |
| **SAMBA II SARS-CoV-2 & COVIDIX IgM & IgG**<br>Positive predictive agreement<br>Negative predictive agreement | 100 (59.0-100)<br>100 (59.0-100) | 100 (71.5-100)<br>66.7 (9.4-99.2) | 100 (54.1-100)<br>81.8 (48.2-97.7) | 100 (85.8-100)<br>85.7(63.7-97.0) |
| **SureScreen IgM & IgG\***<br>Positive predictive agreement<br>Negative predictive agreement | 42.9 (9.9-81.6)<br>100 (54.1-100) | 90.9 (58.7-99.8)<br>66.7 (9.4-99.2) | 100 (54.1-100)<br>100 (69.2-100) | 79.2 (57.8-92.9)<br>94.7 (74.0-99.9) |
| **SAMBA II SARS-CoV-2 & SureScreen IgM & IgG\***<br>Positive predictive agreement<br>Negative predictive agreement | 100 (59.0-100)<br>100 (54.1-100) | 100 (71.5-100)<br>66.7 (9.4-99.2) | 100 (54.1-100)<br>100 (69.2-100) | 100 (85.8-100)<br>94.7(74.0-99.9) |

Three participants had stored samples available for testing at multiple time points in their illness (Figure 2). Two individuals were sampled from early after symptom onset and the third presented three weeks into illness. In the first two cases (Figure 2A-F), we observed an increase in neutralisation activity over time that was mirrored by band intensities on rapid POC antibody testing. As expected IgM bands arose early on with IgG following closely. Of note in patient 1 there was a weakly detectable IgM band by rapid test with no serum neutralisation activity (Figure 2A, B). Over time the band intensity for IgM and IgG increased along with serum neutralisation activity. In the individual presenting 21 days into illness (Figure 2G-I), only IgG was detected with rapid POC antibody testing and as expected band intensity did not increase over the following days.

**Figure 2:**
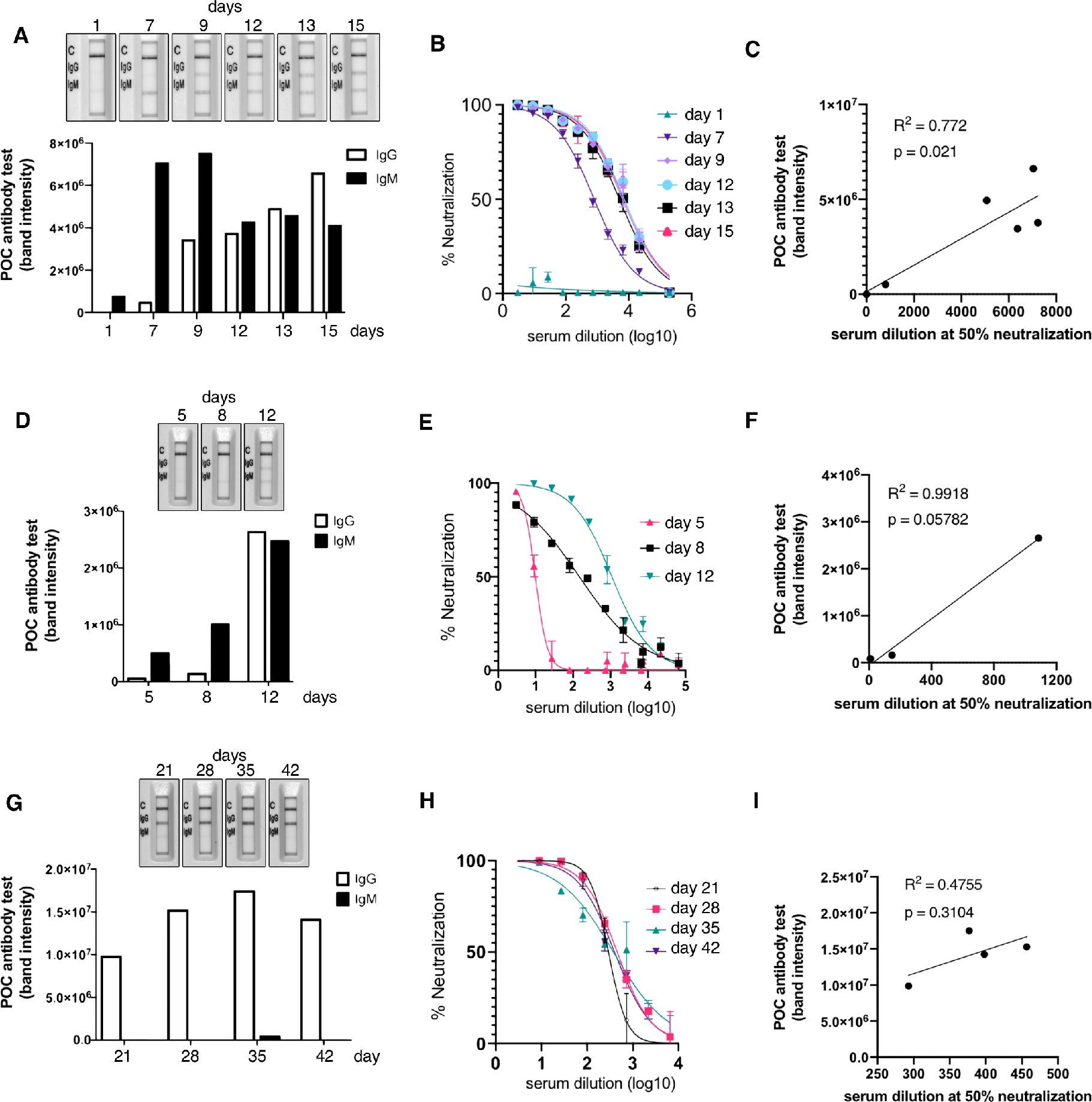
Longitudinal antibody responses detected by rapid lateral flow and neutralisation assays. A, D, G. An immune-chromatographic lateral flow rapid diagnostic test (POC antibody test - COVIDIX SARS-CoV-2 IgM IgG Test) on longitudinal samples in individual patients detecting SARS-CoV-2 IgM and IgG bands. Band intensities were acquired using ChemiDoc MP Imaging System and quantified using Image Lab software. B, E, H. SARS-CoV-2 pseudotyped virus neutralisation assay from longitudinal serum samples in individual patient examples. The assays were performed in duplicate. Error bars represent SEM. C, F, I. Comparison of IgG band intensities from lateral flow rapid diagnostic test with EC50 neutralisation titres from SARS-CoV-2 pseudotyped virus neutralisation assay in individual patients. Correlations were estimated by linear regression analysis.

## Discussion

Here we have shown that POC NAAT testing in combination with antibody detection can improve diagnosis of COVID-19 in moderate to severe suspected cases. Overall positive predictive agreement against the composite reference standard was around 79% for rapid NAAT testing of nose/throat swab samples, reaching 100% with a combined approach of rapid NAAT testing and either of the two POC LFA antibody tests. The expected presence of some false positive antibody rapid results decreased the specificity of the combined approach to 85.7-94.7% overall. As expected, nucleic acid detection in nose/throat samples was highest in those presenting within the first few days (100% in samples taken in the first 4 days after symptom onset). Conversely antibody detection by LFA increased with time since symptom onset with 100% efficacy beyond 9^th^ day post-symptoms.

One study reported that combined lab based RT-PCR with lab based antibody testing could increase sensitivity for COVID-19 diagnosis from 67.1% to 99.4% in hospitalised patients(Zhao et al., 2020). However, in that study this assessment of sensitivity was made using clinical diagnosis. A major strength of this study is the use of an objective reference standard that included NAAT and serum neutralisation - a phenotypic test for functionality of antibodies. This assay was shown to be robust and accurate, using a recently described ELISA method for SARS-CoV-2 IgG detection that is now used globally(Amanat et al., 2020).

Use of antibody tests for COVID-19 diagnosis in hospitals has been limited for a number of reasons. Firstly, we know from SARS-CoV-1 that previous humoral immunity to HCoV OC43 and 229E can elicit a cross-reactive antibody response to N of SARS-CoV-1 in up to 14% of people tested in cross-sectional studies(Woo et al., 2004), and previous exposure to HCoV can rarely elicit a cross-reactive antibody response to the N and S proteins of SARS-CoV-2 (Jaaskelainen et al., 2020; Pickering et al., 2020). Secondly, antibody tests do not achieve the same detection rates as nucleic acid based tests early in infection, as humoral responses take time to develop following viral antigenic stimulation. However, by day 6 post symptom onset detection of IgG to Spike protein has been reported to reach 100% sensitivity (Long et al., 2020) and this is useful in cases with immune mediated inflammatory disease where RT-PCR on respiratory samples is often negative, for example in the recently described Kawasaki-like syndrome named PIMS (paediatric inflammatory multi-system syndrome) (Verdoni et al., 2020).

CT scanning has previously been shown to be highly sensitive(Fang et al., 2020), though few countries have the resources for large scale CT based screening. In our study chest radiographs were significantly more likely to show changes associated with COVID-19, but a quarter of chest radiographs in the confirmed COVID-19 group were normal or indeterminate.

This study had limited numbers of participants, though patients were distributed well by symptom onset and were part of a clinical trial with complete data. We tested stored sera rather than whole blood finger prick, though this was intentional given the caution needed in interpreting antibody tests and potential cross-reactivity of antibodies. Although SARS-CoV-2 ELISA testing of our pre 2020 sera did reveal occasional N and S reactivity to SARS-CoV-2 (Supplementary table 2), these samples were negative on the rapid antibody testing. In light of our data, prospective evaluation on a finger prick sample is now warranted on a larger scale in patients with moderate to severe disease. In the present study, both POC antibody tests were used with serum samples and one of them presented more false positive reactions. It can be predicted that such erroneous results with one but possibly both POC assays will be observed in increased numbers with capillary whole blood samples. For this reason, it would be advisable to perform confirmation either with an alternative POC rapid test or a laboratory based platform. At present we cannot speculate on the diagnostic accuracy of the antibody or NAAT tests in mild disease.

We envisage a deployment approach whereby both test samples, finger prick whole blood and nose/throat swab, are taken at the same time on admission to hospital. The finger prick antibody test result is available within 15 minutes. A positive POC antibody test result as the only positive marker should ideally be confirmed with a second rapid POC test / laboratory IgG/IgM test before movement to a COVID-19 area, or recruitment into a clinical treatment study. The NAAT result remains critical not only to identify early infection but, more importantly to triage infectious patients to be isolated from other patients and be handled with particular care by staff. NAAT is also expected to be more valuable than antibody tests in milder and asymptomatic cases given severity appears to correlate with magnitude of antibody responses (Pickering et al., 2020; Wang et al., 2020a).

Rapid combined tests could be transformative in diagnosis and management of moderate to severe COVID-19 disease requiring hospitalisation, particularly as diverse manifestations of disease emerge.

## Methods

### Cell lines

293T cells were cultured in DMEM complete (DMEM supplemented with 100 U/ml penicillin, 0.1 mg/ml streptomycin, and 10% FCS).

### Pseudotype virus preparation

Viral vectors were prepared by transfection of 293T cells by using Fugene HD transfection reagent (Promega) as follows. Confluent 293T cells were transfected with a mixture of 11ul of Fugene HD, 1µg of pCAGGS_SARS-CoV-2_Spike, 1ug of p8.91 HIV-1 gag-pol expression vector(Gupta et al., 2010; Naldini et al., 1996), and 1.5µg of pCSFLW (expressing the firefly luciferase reporter gene with the HIV-1 packaging signal). Viral supernatant was collected at 48 and 72h after transfection, filtered through 0.45um filter and stored at −80° C. The 50% tissue culture infectious dose (TCID50) of SARS-CoV-2 pseudovirus was determined using Steady-Glo Luciferase assay system (Promega).

### Pseudotype neutralisation assay

Spike pseudotype assays have been shown to have similar characteristics as neutralization testing using fully infectious wild type SARS-CoV-2(Schmidt et al., 2020).Virus neutralization assays were performed on 293T cell transiently transfected with ACE2 and TMPRSS2 using SARS-CoV-2 Spike pseudotyped virus expressing luciferase. Pseudovirus was incubated with serial dilution of heat inactivated human serum samples from COVID-19 suspected individuals in duplicates for 1h at 37° C. Virus and cell only controls were also included. Then, freshly trypsinized 293T ACE2/TMPRSS2 expressing cells were added to each well. Following 48h incubation in a 5% CO2 environment at 37°C, the luminescence was measured using Steady-Glo Luciferase assay system (Promega). The 50% inhibitory dilution (EC50) was defined as the serum dilution at which the relative light units (RLUs) were reduced by 50% compared with the virus control wells (virus + cells) after subtraction of the background RLUs in the control groups with cells only. The EC50 values were calculated with non-linear regression, log (inhibitor) vs. normalized response using GraphPad Prism 8 (GraphPad Software, Inc., San Diego, CA, USA). The neutralisation assay was positive if the serum achieved at least 50% inhibition at 1 in 3 dilution of the SARS-CoV-2 spike protein pseudotyped virus in the neutralisation assay. The neutralisation result was negative if it failed to achieve 50% inhibition at 1 in 3 dilution.

### Enzyme-linked immunosorbent assay (ELISA)

We developed an ELISA targeting the SARS-CoV-2 Spike and N proteins. Trimeric spike protein antigen used in the ELISA assays consists of the complete S protein ectodomain with a C-terminal extension containing a TEV protease cleavage site, a T4 trimerization foldon and a hexa-histidine tag. The S1/S2 cleavage site with amino acid sequence PRRAR was replaced with a single Arginine residue and stabilizing Proline mutants were inserted at positions 986 and 987. Spike protein was expressed and purified from Expi293 cells (Thermo Fisher). N protein consisting of residues 45-365 was initially expressed as a His-TEV-SUMO-fusion. After Ni-NTA purification, the tag was removed by TEV proteolysis and the cleaved tagless protein further purified on Heparin and gel filtration columns.

The ELISAs were in a stepwise process; a positivity screen was followed by endpoint titre as previously described(Amanat et al., 2020). Briefly, 96-well EIA/RIA plates (Corning, Sigma) were coated with PBS or 0.1µg per well of antigen at 4°C overnight. Coating solution was removed, and wells were blocked with 3% skimmed milk prepared in PBS with 0.1% Tween 20 (PBST) at ambient temperature for 1 hour. Previously inactivated serum samples (56°C for 1 hour) were diluted to 1:60 or serially diluted by 3-fold, six times in 1% skimmed milk in PBST. Blocking solution was aspirated and the diluted sera were added to the plates and incubated for 2 hours at ambient temperature. Diluted sera were removed, and plates were washed three times with PBST. Goat anti-human IgG secondary antibody-Peroxidase (Fc-specific, Sigma) prepared at 1:3,000 in PBST was added and plates were incubated for 1 hour at ambient temperature. Plates were washed three times with PBST. ELISAs were developed using 3,5,3′,5′-tetramethylbenzidine (TMB, ThermoScientific); reactions were stopped after 10 minutes using 0.16M Sulfuric acid. The optical density at 450 nm (OD450) was measured using a Spectramax i3 plate reader. The absorbance values for each sample were determined by subtracting OD values from uncoated wells. All data analyses were performed using Prism 8 version 8.4.2 (GraphPad). An OD cut off of 0.3 was used to define a positive IgG response to full length Spike protein.

### COVIDIX 2019 SARS-CoV-2 IgG/IgM Test (COVIDIX Healthcare, Cambridge, UK)

This colloidal-gold lateral flow immunoassay is designed to detect IgG and IgM to SARS-CoV-2. The test is CE marked. It was used according to the manufacturer’s instructions. 10µl of serum was added to the test well followed by 2 drops of the manufacturer’s proprietary buffer. In order to rule out cross reactivity of this test with seasonal coronavirus antibodies we tested 19 stored specimens from before 2020, some of which had N and S protein SARS-CoV-2 cross reactivity (Supplementary table 2). For quantification of IgG and IgM band density in COVIDIX 2019 nCoV IgG/IgM Test, high resolution images of completed POC antibody test cassettes were acquired using ChemiDoc MP Imaging System (Bio-Rad) at 20min post-addition of the human serum. Band intensities were analysed using Image Lab software (Bio-Rad).

### SureScreen SARS-CoV-2 IgG/IgM Test (SureScreen Diagnostics Ltd, Derby, UK)

This colloidal-gold lateral flow immunoassay is designed to detect IgG and IgM to SARS-CoV-2. It was used according to the manufacturer’s instructions. The test has been CE marked and previously validated against a large panel of negative historical controls and in serum from confirmed PCR positive COVID-19 cases(Pickering et al., 2020). 10µl of serum was added to the test well followed by 2 drops of the manufacturer’s proprietary buffer.

### Participants

The study participants were part of the COVIDx trial(Collier et al., 2020), a prospective analytical study which compared SAMBA II SARS-CoV-2 point of care test to the standard laboratory RT-PCR test for the detection of SARS-CoV-2 in participants admitted to Cambridge University Hospitals NHS Foundation Trust (CUH) with a possible diagnosis of COVID-19. Consecutive participants were recruited during 12-hour day shifts over a duration of 4 weeks from the 6^th^ of April 2020 to the 2^nd^ of May 2020. We recruited adults (>16 years old) presenting to the emergency department or acute medical assessment unit as a possible case of COVID-19 infection. This included any adult requiring hospital admission and who was symptomatic of SARS-CoV-2 infection, demonstrated by clinical or radiological findings. (Collier et al., 2020). 48 participants who had available stored sera were included in this sub-study and underwent further antibody testing. The laboratory standard RT-PCR test, developed by public health England (PHE), targeting the RdRp gene was performed on a combined nose/throat swab in parallel. This test has an estimated limit of detection of 320 copies/ml. SAMBA II SARS-CoV-2 testing was performed on a combined nose/throat swab collected by dry sterile swab and inactivated in a proprietary buffer at point of sampling. SAMBA II SARS-CoV-2 targets 2 genes-Orf1 and the N genes and uses nucleic acid sequence based amplification to detect SARS-CoV-2 RNA, with limit of detection of 250 copies/ml.

### Assessment of neutralisation assay performance

Four assays detecting IgG to COVID-19 were utilised in this study. 38 of the 45 samples were identified as concordant with at least three of the four assays and considered confirmed either negative or positive. Against this group of samples validated for content of COVID-19 IgG, each individual assay was assessed. Neutralisation, ELISA, SureScreen and COVIDIX assays gave a correct result in 100%, 97.4%, 92.1% and 86.8%, respectively, justifying the choice of the neutralisation assay as standard.

### Analyses

The performance of SAMBA II SARS-CoV-2 test and COVIDIX SARS-CoV-2 IgG/IgM Test or SureScreen SARS-CoV-2 IgG/IgM Test for diagnosing COVID-19 were calculated alone and then in combination along with binomial 95% confidence intervals (CI). A composite reference standard was used - standard lab RT-PCR and a neutralisation assay. Descriptive analyses of clinical and demographic data are presented as median and interquartile range (IQR) when continuous and as frequency and proportion (%) when categorical. The differences in continuous and categorical data were tested using Wilcoxon rank sum and Chi-square test respectively. Statistical analysis were conducted using Stata (version 13), with additional plots generated using GraphPad Prism.

## Data Availability

Data are available on request from the corresponding author

## Acknowledgements

we would like to thank Jakub Luptak, Martin Besser, Rainer Doffinger, Helen Lee, Gabriel Hawthorn and Sara Lear. pCAGGS_SARS-CoV-2_Spike was obtained by CFAR, NIBSC, thanks to the donation of Dr Emma Bentley.

## Ethical approval

COVIDx (NCT04326387) was approved by the East of England - Essex Research Ethics Committee (REC ref: 20/EE/0109). Serum samples were obtained from patients attending Addenbrooke’s Hospital with a suspected or confirmed diagnosis of COVID19. Ethical approval was obtained from the East of England – Cambridge Central Research Ethics Committee (REC ref 17/EE/0025).

---

**Supplementary Figure 1:**
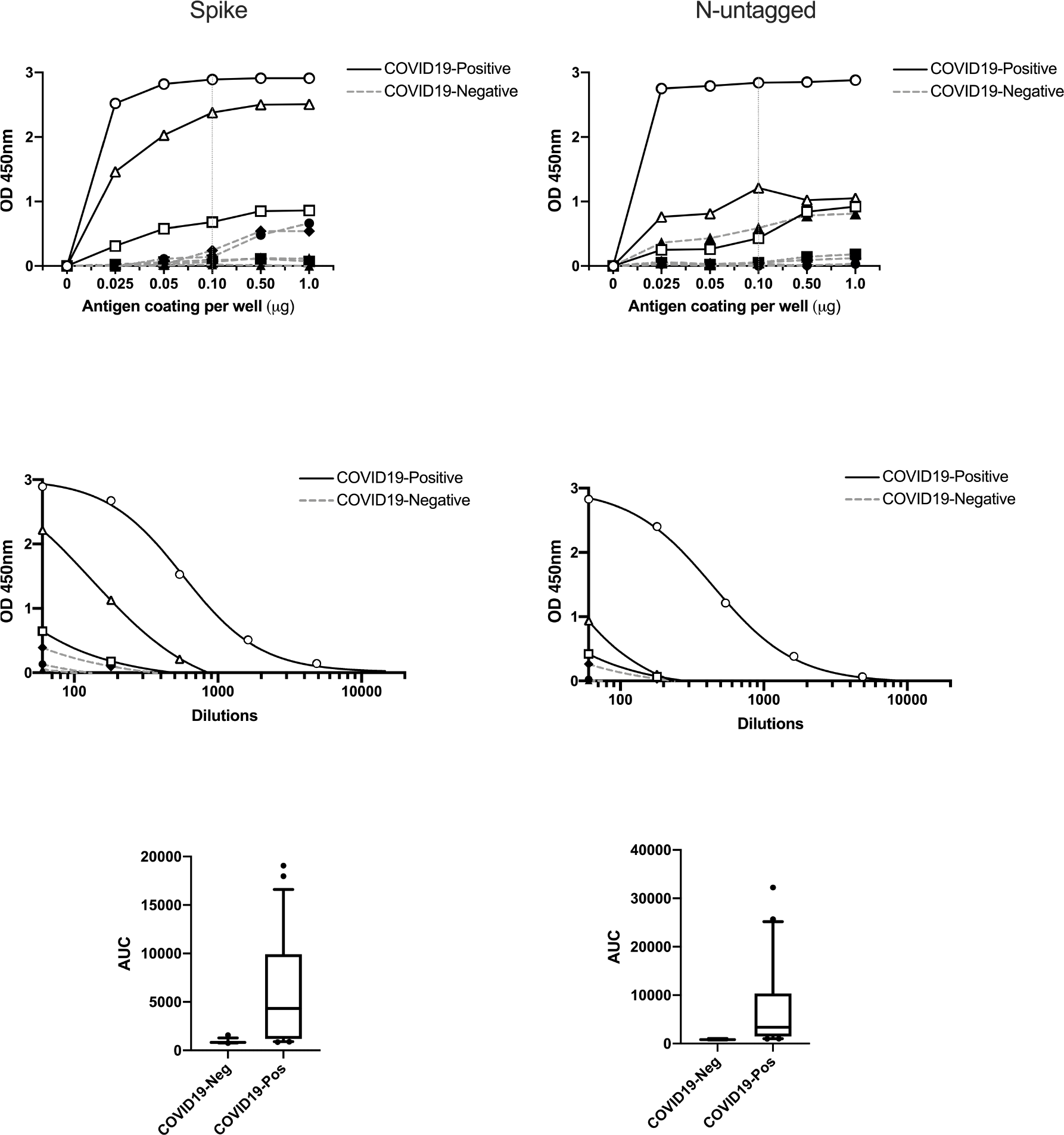
Establishment of serological assay to determine positivity and endpoint titre against human SARS CoV-2. Residual stored serum samples from PCR positive and negative patient cohort were screened for reactivity against full-length spike and N-proteins. A) To determine the appropriate concentration of antigen used for plate coating, 0, 0.025, 0.05, 0.1, 0.5 and 1.0 1mg antigen per well was coated and reactivity of known seropositive and seronegative serum samples were examined. B) Subsequently, end-point titrations were performed using 0.1mg per well spike and N antigen coating. C. The area under the curve (AUC) was calculated for every sample using end point titrations against spike (n=76) and N protein (n=64), and the mean and the 95% confidence intervals are shown for all PCR positive and negative samples. OD: optical density (nanometers)

**Supplementary Figure 2:**
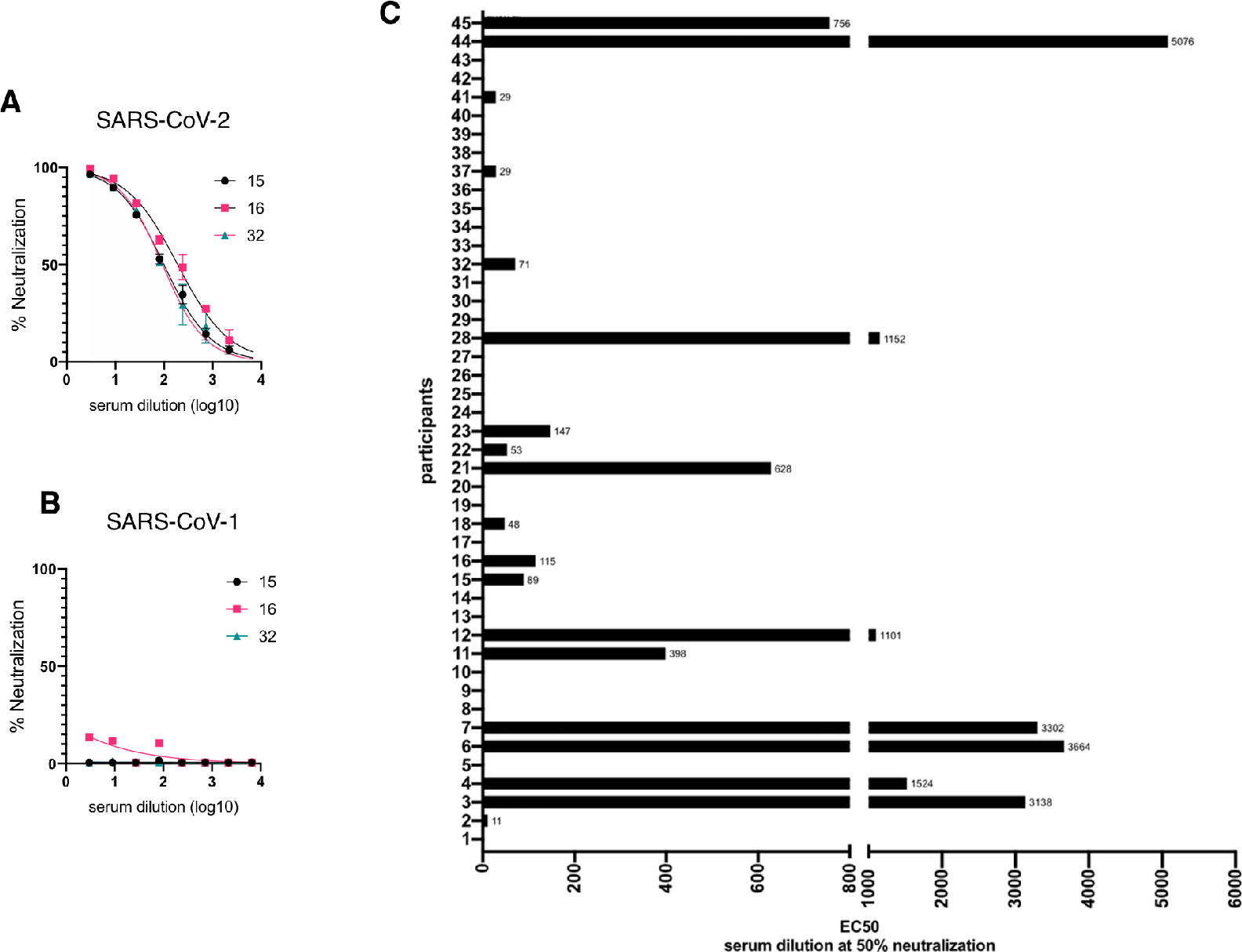
Specificity of antibody neutralising response against SARS-CoV-2 and CoV-1. SARS-CoV-2 **(A)** or SARS-CoV-1 **(B)** Spike protein pseudotyped viral particles were incubated with serial dilutions of heat inactivated human serum samples from Covid-19 suspected individuals (#15,16,32) in duplicates for 1h at 37° C. 293T ACE2/TMPRSS2 expressing cells were added to each well. Following 48h incubation in a 5% CO2 environment at 37°C, the luminescence was measured using Steady-Glo Luciferase assay system (Promega). Percentage of neutralisation was calculated with non-linear regression, log (inhibitor) vs. normalized response using GraphPad Prism 8 (GraphPad Software, Inc., San Diego, CA, USA). **(C)** The 50% inhibitory dilution (EC50) was defined as the serum dilution at which the relative light units (RLUs) were reduced by 50% compared with the virus control wells (virus + cells) after subtraction of the background RLUs in the control groups with cells only. The EC50 values were calculated with non-linear regression, log (inhibitor) vs. normalized response using GraphPad Prism 8 (GraphPad Software, Inc., San Diego, CA, USA).

**Supplementary Figure 3:**
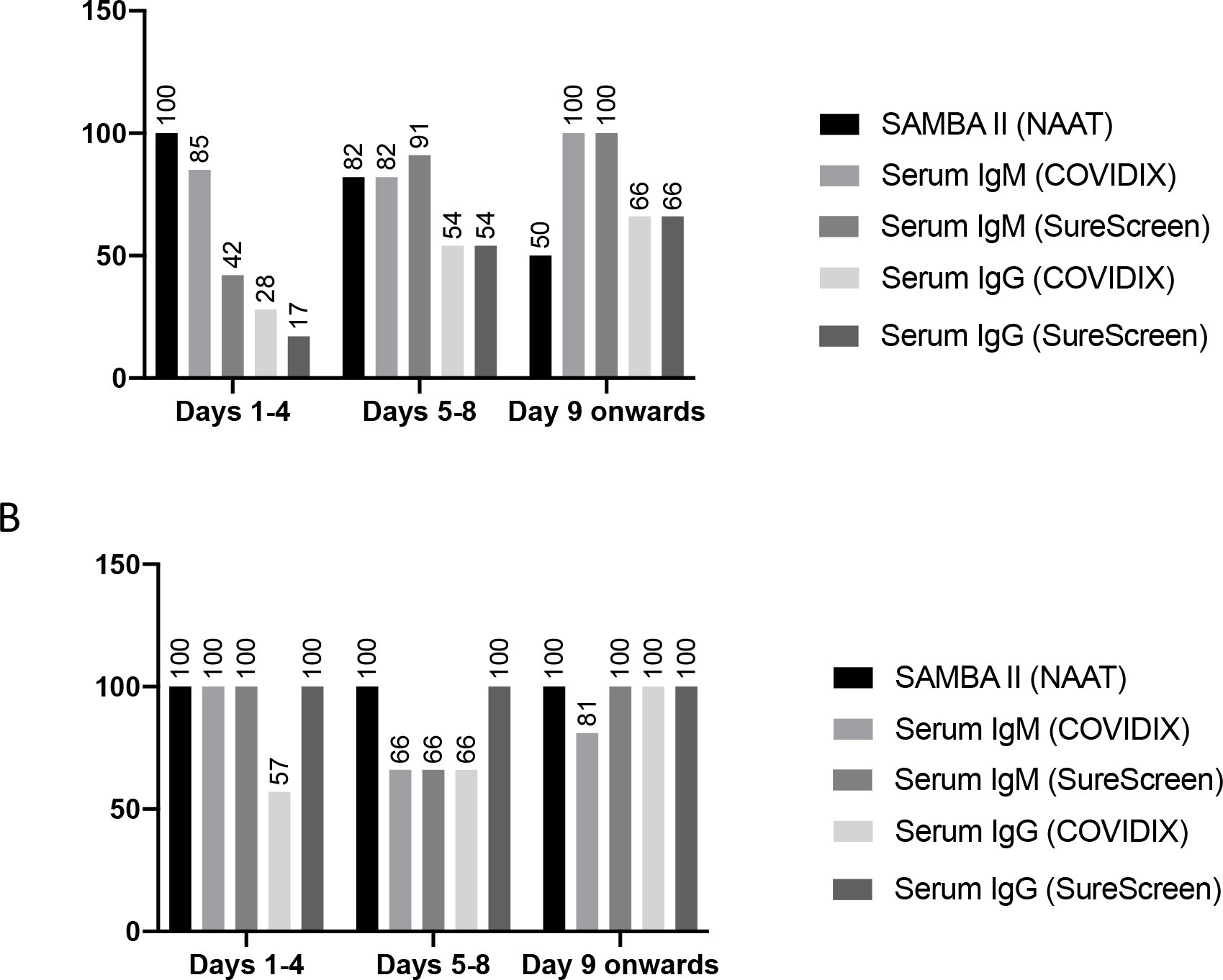
Results of assays by time since onset of symptoms. (A) % of positive tests in individuals classified as COVID-19 positive by composite reference (B) % of negative tests in individuals classified as COVID-19 negative by composite reference.

**Supplementary Table 1:** clinical details of participants with false positive combined rapid testing (due to false positive rapid IgM/ IgG result).

| **Clinical features** | **#20** | **#35** | **#40** |
| --- | --- | --- | --- |
| Radiology | Normal | Normal | Pulmonary embolus |
| Oxygen saturations (%) | 95 | 88 | 97 |
| PaO<sub>2</sub>/FiO<sub>2</sub> | 42.9 | 41.9 | 32.4 |
| Temperature ( °C) | 37.0 | 37.9 | 36.9 |
| Respiratory rate (breaths/min) | 17 | 24 | 18 |
| Lymphocyte count (x10<sup>9</sup> /L) | 3.3 | 1.7 | 1.2 |
| C-reactive protein (mg/L) | 4 | 35 | 135 |

**Supplementary Table 2:** Pre 2020 sera testing: ELISA optical density values for full length SARS-CoV-2 Spike (FL), Spike receptor binding domain (RBD), nucleocapsid (N), and result on testing with COVIDIX SARS-CoV-2 IgM/IgG test. Positive (from confirmed positive) and negative (pooled human sera from pre 2020) control values are given.

| Sample no | FL | RBD | N | COVIDIX IgM/IgG result |
| --- | --- | --- | --- | --- |
| 1 | 0.95735 | 0.20455 | 0.5343 | Negative |
| 2 | 0.1217 | 0.1008 | 0.0746 | Negative |
| 3 | 0.2680 | 0.1300 | 0.1285 | Negative |
| 4 | 0.2511 | 0.0837 | 0.07445 | Negative |
| 5 | 0.10625 | 0.06625 | 0.4722 | Negative |
| 6 | 0.1561 | 0.08655 | 0.0927 | Negative |
| 7 | 1.12375 | 0.05785 | 0.40535 | Negative |
| 8 | 0.1432 | 0.0888 | 0.5842 | Negative |
| 9 | 0.49075 | 0.06505 | 0.32445 | Negative |
| 10 | 0.16075 | 0.03625 | 0.13485 | Negative |
| 11 | 0.08205 | 0.0504 | 0.07485 | Negative |
| 12 | 0.1956 | 0.23025 | 0.1748 | Negative |
| 13 | 0.1482 | 0.07115 | 0.05645 | Negative |
| 14 | 0.16075 | 0.078 | 1.00845 | Negative |
| 15 | 0.18015 | 0.09845 | 0.7598 | Negative |
| 16 | 0.26335 | 0.0693 | 0.38865 | Negative |
| 17 | 0.1864 | 0.18905 | 0.35065 | Negative |
| 18 | 0.1265 | 0.3684 | 0.18025 | Negative |
| 19 | 0.08425 | 0.06555 | 0.1378 | Negative |
| Negative | 0.297 | 0.054 | 0.387 |  |
| Positive | 2.704 | 2.150 | 2.337 |  |

